# Spectrum and Immunovirological Determinants of Tumours Among People Living with HIV in Uganda: A Retrospective Cohort Study from a Specialised HIV Centre, 2017–2026

**DOI:** 10.64898/2026.04.30.26352124

**Authors:** Joseph Mwaka, Conrad Sserunjogi, Jessica Natalayi

## Abstract

**Background:** Sub-Saharan Africa carries a disproportionate burden of HIV-associated malignancies, yet the contemporary tumour spectrum in the dolutegravir (DTG) era and the immunovirological context at cancer diagnosis remain incompletely characterised in East African routine HIV-care settings. We sought to describe the spectrum, malignant fraction, and immunovirological determinants of neoplasms diagnosed at a specialised HIV centre in Kampala, Uganda.

**Methods:** We conducted a retrospective cohort analysis of 220 tumour records from 217 unique people living with HIV (PLHIV) registered at Mildmay Hospital between November 2017 and January 2026. Tumours were classified by ICD-10 disease group and malignancy status (benign/malignant). Bivariate associations with malignancy status were tested using χ^2^ or Fisher exact tests for categorical variables and Mann–Whitney U tests for continuous variables (α = 0.05). Crude odds ratios (OR) with 95% confidence intervals (CI) were computed for selected dichotomous comparisons.

**Results:** The cohort had a median age of 47 years (IQR 39–54) and was 58.2% female. Overall, 138/220 tumours (62.7%) were malignant. Kaposi sarcoma (KS) was the most frequent malignancy (n=65; 47.1% of all malignant cases), followed by haematopoietic malignancies (n=32; 23.2%) and malignant gynaecological tumours (n=10; 7.2%). Among 156 patients with viral load data, 134 (85.9%) were virally suppressed at tumour diagnosis, yet 70/134 (52.2%) of those suppressed patients had a malignant tumour. Advanced WHO clinical stage (III/IV) at ART initiation was strongly associated with malignant diagnosis (39/86 [45.3%] versus 9/79 [11.4%]; OR 3.53, 95% CI 1.59–7.84; *p*=0.002). Median CD4 at ART initiation was markedly lower in malignant compared to benign tumour patients (147 vs 476 cells/µL; p<0.001), and median ART duration was substantially shorter (37 vs 102 months; p<0.001). KS patients had significantly lower CD4 (136 vs 269 cells/µL; *p*=0.002) and shorter ART duration (4 vs 86 months; p<0.001) than patients with other malignancies. Malignancy did not differ across DTG-, EFV-, and PI-based regimens *(p*=0.992).

**Conclusions:** HIV-associated tumours in this Ugandan cohort remain predominantly malignant and dominated by KS, even against a backdrop of high viral suppression rates. The clustering of malignancy among patients with low pre-ART CD4 counts and advanced WHO stage points to incomplete immune reconstitution, persistent oncoviral co-infection, and residual inflammation as mechanistic targets. These data establish the scientific platform for a prospective HIV-oncology cohort at Mildmay Hospital and Mildmay Research Centre and inform context-appropriate cancer screening and surveillance priorities.

## 1. Introduction

The relationship between HIV infection and cancer has been fundamentally reshaped by the global scale-up of combination antiretroviral therapy (ART). In the pre-ART era, the three AIDS-defining cancers: Kaposi sarcoma (KS), non-Hodgkin lymphoma (NHL), and invasive cervical cancer dominated the malignancy landscape of people living with HIV (PLHIV) and were direct manifestations of advanced immunosuppression [1,2]. With the advent of sustained viral suppression, the relative burden of AIDS-defining cancers declined in high-income settings, while non-AIDS-defining cancers (NADCs) including hepatocellular, lung, anal, and colorectal carcinomas rose, driven by an ageing PLHIV population, chronic inflammation, and persistent oncogenic co-infections such as human papillomavirus (HPV), hepatitis B and C viruses, Epstein– Barr virus (EBV), and human herpesvirus-8 (HHV-8) [3–6].

In sub-Saharan Africa, this epidemiological transition is incomplete and geographically uneven. KS remains the most frequently reported cancer among PLHIV across East Africa, reflecting an exceptionally high regional seroprevalence of HHV-8 (often exceeding 40% in adult populations) and persistent immune reconstitution gaps that follow late ART initiation [7–10]. Uganda carries one of the highest age-standardised KS incidence rates globally. National data from the Kampala Cancer Registry (Africa’s longest-running population-based cancer registry) document a continuing predominance of infection-associated malignancies among PLHIV, even in the contemporary treatment era [11–14].

Despite this burden, the cancer spectrum among Ugandan PLHIV in the era of dolutegravir (DTG)-based first-line therapy adopted as Uganda’s preferred ART regimen following WHO 2018–2019 guidelines [15,16] has not been systematically characterised within specialised HIV clinical settings. Most published evidence derives either from population-based cancer registries that lack ART history and virological data, or from oncology units that under-record HIV exposure [13,17,18]. Consequently, the joint immunological-virological-oncological profile of tumours diagnosed in routine HIV care including the contribution of co-occurring benign neoplasms remains poorly defined.

Three specific evidence gaps motivate this study. First, the proportion of tumours that are malignant versus benign in HIV clinic populations is rarely reported, yet it is operationally important: benign tumours such as uterine leiomyomas and benign prostatic hyperplasia consume substantial diagnostic and surgical capacity and may be modulated by chronic ART exposure [19,20]. Second, the immunovirological context at cancer diagnosis; viral load, CD4 nadir, WHO stage at ART initiation, ART duration, and ART backbone are needed to identify subgroups at greatest residual risk in the era of high population-level viral suppression. Third, with the growing push towards integrated HIV–non-communicable disease (NCD) services, data anchored in HIV clinical settings are needed to inform context-appropriate cancer screening and referral pathways [21,22].

Mildmay Hospital is a specialised HIV hospital with embedded Research Centre (Mildmay Research Centre) located in Lweza, Kampala, Uganda, with longitudinal electronic records spanning the policy transition from efavirenz (EFV)-based to DTG-based first-line ART. We leveraged nearly a decade of these records to provide the first immunovirologically contextualised description of tumours diagnosed among PLHIV at a Ugandan specialised HIV Centre. The findings establish the epidemiological foundation for a prospective HIV-oncology cohort and directly inform site-level surveillance, diagnostic, and referral priorities.

## 2. Methods

### 2.1 Study design and setting

This was a retrospective observational cohort study based on routinely collected electronic clinical records at Mildmay Hospital, Kampala, Uganda. Mildmay is a specialised HIV Centre providing comprehensive HIV testing, ART initiation, virological monitoring, and management of HIV-related comorbidities, including referral pathways for malignancies. The Mildmay Research Centre which is the Research arm of Mildmay Uganda is located within the same compass with Mildmay Hospital. The Centre’s electronic data system has captured ICD-10-coded diagnoses since 2017. There are approximately 14,500 patients in active care and over 17,000 patient records (as of January 2026).

### 2.2 Study population and case definition

All PLHIV with at least one ICD-10 tumour diagnosis recorded at Hospital Electronic medical registry between 1 November 2017 and 29 January 2026 were eligible. The primary analytical unit was the tumour record, irrespective of the modality by which it was diagnosed (clinical, radiological, pathological). Three patients contributed two distinct primary tumour records each on different dates; this is disclosed in the Methods and addressed in the Limitations. Tumours were classified as malignant or benign based on ICD-10 coding and aggregated into five clinically meaningful groups: Kaposi sarcoma, haematopoietic malignancies, gynaecological tumours, prostate-related tumours, and other solid tumours.

### 2.3 Variables and definitions

Demographic variables: age at tumour diagnosis (years; categorised <30, 30–44, 45–59, ≥60) and sex. Immunological variables: CD4 cell count at ART initiation (cells/µL; categorised <200, 200– 499, ≥500) and WHO clinical stage at ART initiation (I, II, III, IV). Virological variables: plasma HIV RNA at tumour diagnosis, dichotomised as suppressed (<1,000 copies/mL) or unsuppressed (≥1,000 copies/mL) per the prevailing Uganda Ministry of Health threshold [23]. Treatment variables: ART backbone at tumour diagnosis, ART regimen category (DTG-based; EFV-based; other, predominantly PI-based or nevirapine-based), and ART duration from ART initiation to tumour diagnosis in months (categorised ≤12, 13–60, 61–120, >120 months).

### 2.4 Data sources and quality

De-identified records were extracted from the Mildmay Hospital electronic clinical database. ICD-10 codes were verified against the WHO ICD-10 reference [24]. Data quality checks included range validation for age, ART duration, and CD4 counts; cross-checks of WHO stage codes against textual entries; and reconciliation of duplicate patient identifiers. Missingness was quantified for each variable and analyses performed on complete cases for each variable.

### 2.5 Statistical analysis

Continuous variables were summarised as median (IQR) given right-skewed distributions confirmed by the Shapiro–Wilk test; categorical variables as frequencies and percentages. Bivariate associations between candidate exposures and malignancy status were tested with Pearson χ^2^ or Fisher exact tests (the latter when any expected cell count was <5) for categorical variables, and Mann–Whitney U tests for continuous variables. Two-sided p-values <0.05 were considered statistically significant. Crude ORs with 95% CIs were computed where 2×2 tables permitted. No multivariable regression was performed at this descriptive stage; all results are hypothesis-generating. All analyses were conducted in R (version 4.3) using base statistics and the tidyverse framework; results were independently verified by re-running on the source dataset.

### 2.6 Ethical considerations

The study used routinely collected, de-identified clinical data. Ethical approval was obtained from the Mildmay Research Ethics Committee (ref: MUREC-2025-808). Given the retrospective nature and use of de-identified records, the requirement for individual informed consent was waived. The study adhered to the Declaration of Helsinki [25] and STROBE reporting guidelines for observational studies [26].

## 3. Results

### 3.1 Cohort characteristics

A total of 220 tumour records from 217 unique PLHIV met inclusion criteria; three patients contributed two distinct primary tumour records each. The cohort had a median age of 47 years (IQR 39–54; range 7–87), and 128 (58.2%) participants were female. Most tumour records were registered between 2017 and 2019 (n=129; 58.6%), with a peak in 2018 (n=73) partly attributable to a Centre-wide data quality improvement programme, followed by a steady annual flow of 10– 19 new records thereafter through 2025. Cohort characteristics with missingness patterns are summarised in Table 1.

**Table 1.** Baseline characteristics of the cohort (N = 220 tumour records from 217 PLHIV).

| Characteristic | n / Median | % / IQR | Missing, n (%) |
| --- | --- | --- | --- |
| **Age, years, median (IQR)** | 47 | 39–54 | 0 (0) |
| <30 | 17 | 7.7 | — |
| 30–44 | 84 | 38.2 | — |
| 45–59 | 87 | 39.5 | — |
| ≥60 | 32 | 14.5 | — |
| **Sex** |  |  | 0 (0) |
| Female | 128 | 58.2 | — |
| Male | 92 | 41.8 | — |
| **WHO clinical stage at ART initiation (n = 204)** |  |  | 16 (7.3) |
| Stage I | 101 | 49.5 | — |
| Stage II | 55 | 27.0 | — |
| Stage III | 23 | 11.3 | — |
| Stage IV | 25 | 12.3 | — |
| **CD4 count at ART initiation, cells/μL, median (IQR)** | 171 | 87–423 | 127 (57.7) |
| <200 cells/μL | 52 | 55.9 | — |
| 200–499 cells/μL | 22 | 23.7 | — |
| ≥500 cells/μL | 19 | 20.4 | — |
| **ART duration before tumour diagnosis, months, median (IQR)** | 62 | 6–130 | 9 (4.1) |
| ≤12 months | 62 | 29.4 | — |
| 13–60 months | 41 | 19.4 | — |
| 61–120 months | 44 | 20.9 | — |
| >120 months | 64 | 30.3 | — |
| **ART regimen category at tumour diagnosis (n = 178)** |  |  | 42 (19.1) |
| DTG-based | 77 | 43.3 | — |
| EFV-based | 50 | 28.1 | — |
| Other (PI-based / NVP-based) | 51 | 28.7 | — |
| **HIV RNA status at tumour diagnosis (n = 156)** |  |  | 64 (29.1) |
| Suppressed (<1,000 copies/mL) | 134 | 85.9 | — |
| Unsuppressed (≥1,000 copies/mL) | 22 | 14.1 | — |
| **Tumour status** |  |  | 0 (0) |
| Malignant | 138 | 62.7 | — |
| Benign | 82 | 37.3 | — |

ART, antiretroviral therapy; CD4, cluster of differentiation 4; DTG, dolutegravir; EFV, efavirenz; IQR, interquartile range; NVP, nevirapine; PI, protease inhibitor.

Non-trivial missingness reflected routine data realities: CD4 count at ART initiation was unavailable for 127/220 (57.7%) records, HIV RNA at tumour diagnosis was missing for 64/220 (29.1%), ART regimen was unrecorded in 42/220 (19.1%), and WHO clinical stage was unknown in 16/220 (7.3%). The high rate of missing CD4 data likely reflects that a substantial proportion of patients were enrolled in the electronic system only after initial lab values had been obtained elsewhere, and that patients on stable long-term suppressive ART are monitored less intensively.

### 3.2 Tumour spectrum and malignant fraction

Of 220 tumours, 138 (62.7%) were malignant and 82 (37.3%) benign. Five disease groups accounted for the full cohort (Table 2). KS was the largest single disease group (n=65, 29.5% of all tumours; 100% malignant), with cutaneous KS the dominant subtype (n=47, 72.3%). Gynaecological tumours (n=55, 25.0%) were predominantly benign uterine leiomyomas (n=45, 81.8%), with 10 malignant cervical and other gynaecological cancers. The ‘other solid tumours’ group (n=53, 24.1%) was approximately evenly divided between benign (n=25) and malignant (n=28) tumours, encompassing hepatocellular carcinoma, brain tumours, colorectal, and skin cancers. Haematopoietic malignancies (n=32, 14.5%; 100% malignant) were dominated by myelodysplastic syndromes (n=10), lymphomas (n=12 combined Hodgkin and non-Hodgkin), and multiple myeloma (n=4). Prostate-related tumours (n=15, 6.8%) were predominantly benign prostatic hyperplasia (n=12), with three prostate cancers. Combined AIDS-defining cancers (KS, NHL, Hodgkin lymphoma, and invasive cervical cancer) represented 86/220 (39.1%) of the total cohort and 86/138 (62.3%) of all malignant tumours.

**Table 2.** Tumour spectrum by disease group and malignancy status, with leading ICD-10 diagnoses (N = 220).

| Disease group / Leading ICD-10 diagnoses | Total n (%) | Malignant n (%) | Benign n (%) | Leading subtypes |
| --- | --- | --- | --- | --- |
| **Kaposi sarcoma** | 65 (29.5) | 65 (100.0) | 0 (0.0) |  |
| Kaposi sarcoma of skin | 47 | 47 | — | HHV-8-associated; ICD-10: C46.0 |
| Kaposi sarcoma of soft tissue | 7 | 7 | — | ICD-10: C46.1 |
| Kaposi sarcoma of palate | 6 | 6 | — | ICD-10: C46.2 |
| Kaposi sarcoma, other/unspecified | 5 | 5 | — | ICD-10: C46.7/C46.9 |
| **Gynaecological tumours** | 55 (25.0) | 10 (18.2) | 45 (81.8) |  |
| Uterine leiomyoma (all subtypes) | 45 | — | 45 | ICD-10: D25.x; benign |
| Cervical malignancy (exo-/endocervix) | 10 | 10 | — | ICD-10: C53.x; AIDS-defining |
| **Other solid tumours** | 53 (24.1) | 28 (52.8) | 25 (47.2) |  |
| Benign lipomatous neoplasm (all sites) | 12 | — | 12 | ICD-10: D17.x |
| Squamous/basal cell carcinoma of skin | 5 | 5 | — | ICD-10: C44.x |
| Hepatic and gastrointestinal cancers | 5 | 5 | — | ICD-10: C22.x, C18–C20 |
| Other (brain, breast, bladder, etc.) | 31 | 18 | 13 | Mixed NADCs |
| **Haematopoietic malignancies** | 32 (14.5) | 32 (100.0) | 0 (0.0) |  |
| Hodgkin and non-Hodgkin lymphoma | 12 | 12 | — | ICD-10: C81–C86; NHL AIDS-defining |
| Myelodysplastic syndromes/refractory anaemia | 10 | 10 | — | ICD-10: D46.x |
| Multiple myeloma | 4 | 4 | — | ICD-10: C90.0 |
| Other (mycosis fungoides, leukaemia, polycythaemia) | 6 | 6 | — | ICD-10: C84, C91–C95, D45 |
| **Prostate-related tumours** | 15 (6.8) | 3 (20.0) | 12 (80.0) |  |
| Benign prostatic neoplasm / hyperplasia | 12 | — | 12 | ICD-10: D29.1 / N40 |
| Prostate cancer | 3 | 3 | — | ICD-10: C61; NADC |
| **TOTAL** | **220 (100)** | **138 (62.7)** | **82 (37.3)** |  |

ICD-10, International Classification of Diseases, 10th revision; NADC, non-AIDS-defining cancer; KS, Kaposi sarcoma; HHV-8, human herpesvirus-8.

### 3.3 Sex-stratified tumour pattern

The tumour pattern differed substantially by sex. Among 128 women, gynaecological tumours were the largest single group (55/128, 43.0%), most of which were benign uterine leiomyomas (45/55, 81.8%). KS accounted for 24/128 (18.8%) of tumours in women. Among 92 men, KS dominated (41/92, 44.6%; all malignant), followed by other solid tumours (19/92, 20.7%) and haematopoietic malignancies (17/92, 18.5%). Overall, men had a significantly higher malignant tumour fraction than women (73/92 [79.3%] vs 65/128 [50.8%]; χ^2^ p<0.001; OR male vs female 3.72, 95% CI 2.02–6.87), driven primarily by the male predominance of KS and the high frequency of benign uterine fibroids in women.

### 3.4 Immunovirological profile and ART exposure at tumour diagnosis

Bivariate associations with malignancy status are presented in Table 3 and illustrated in Figures 1–3.

**Table 3.**
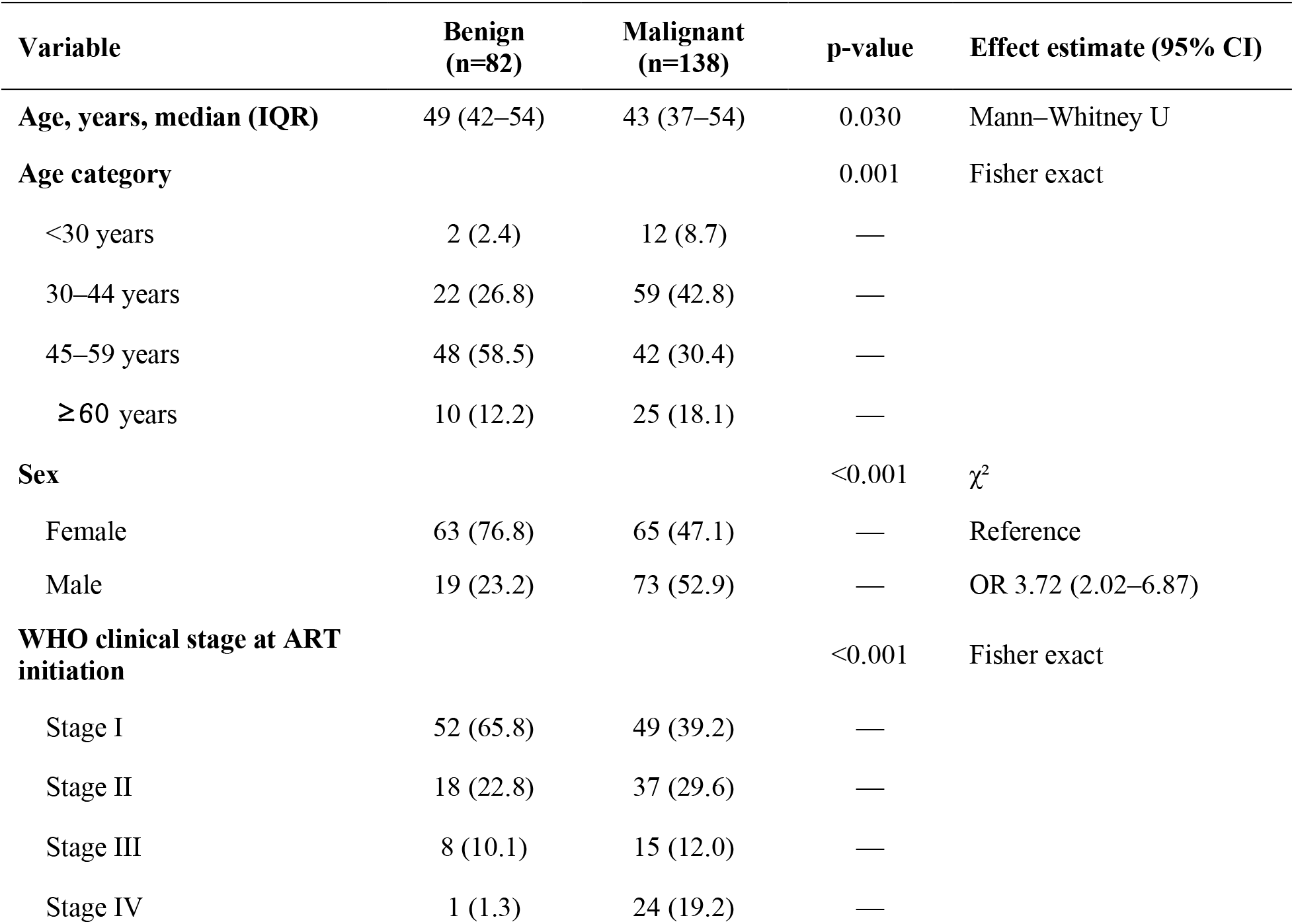
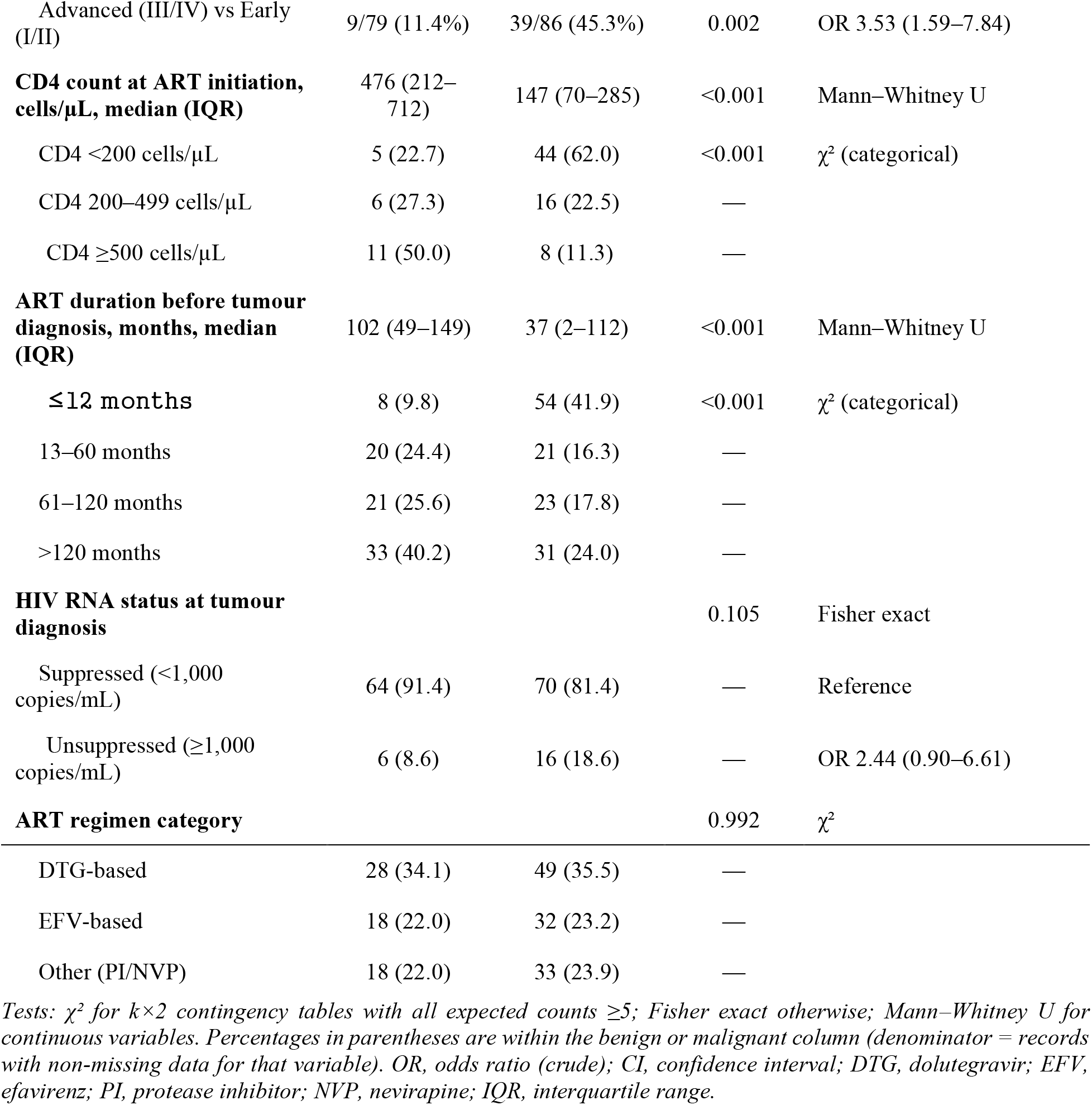
Bivariate associations between patient and treatment characteristics and malignancy status (N = 220 tumour records).

**Figure 1.**
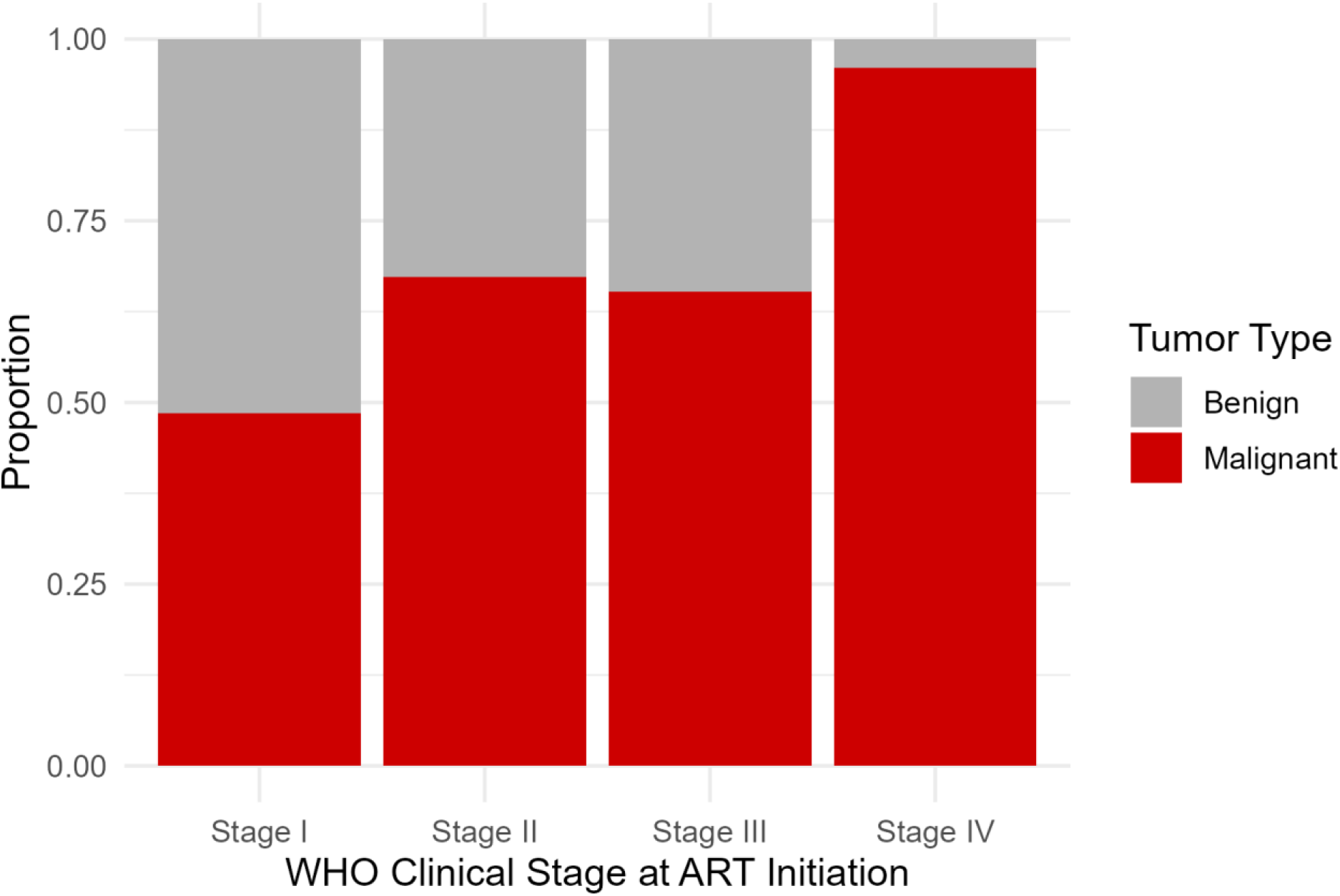
Malignant fraction by WHO clinical stage at ART initiation. Stacked bar chart showing the proportion of malignant (red) versus benign (grey) tumours within each WHO clinical stage group at ART initiation (N = 204 records with known WHO stage). The near-complete malignant enrichment at Stage IV (96%) is visually prominent.

**Figure 2.**
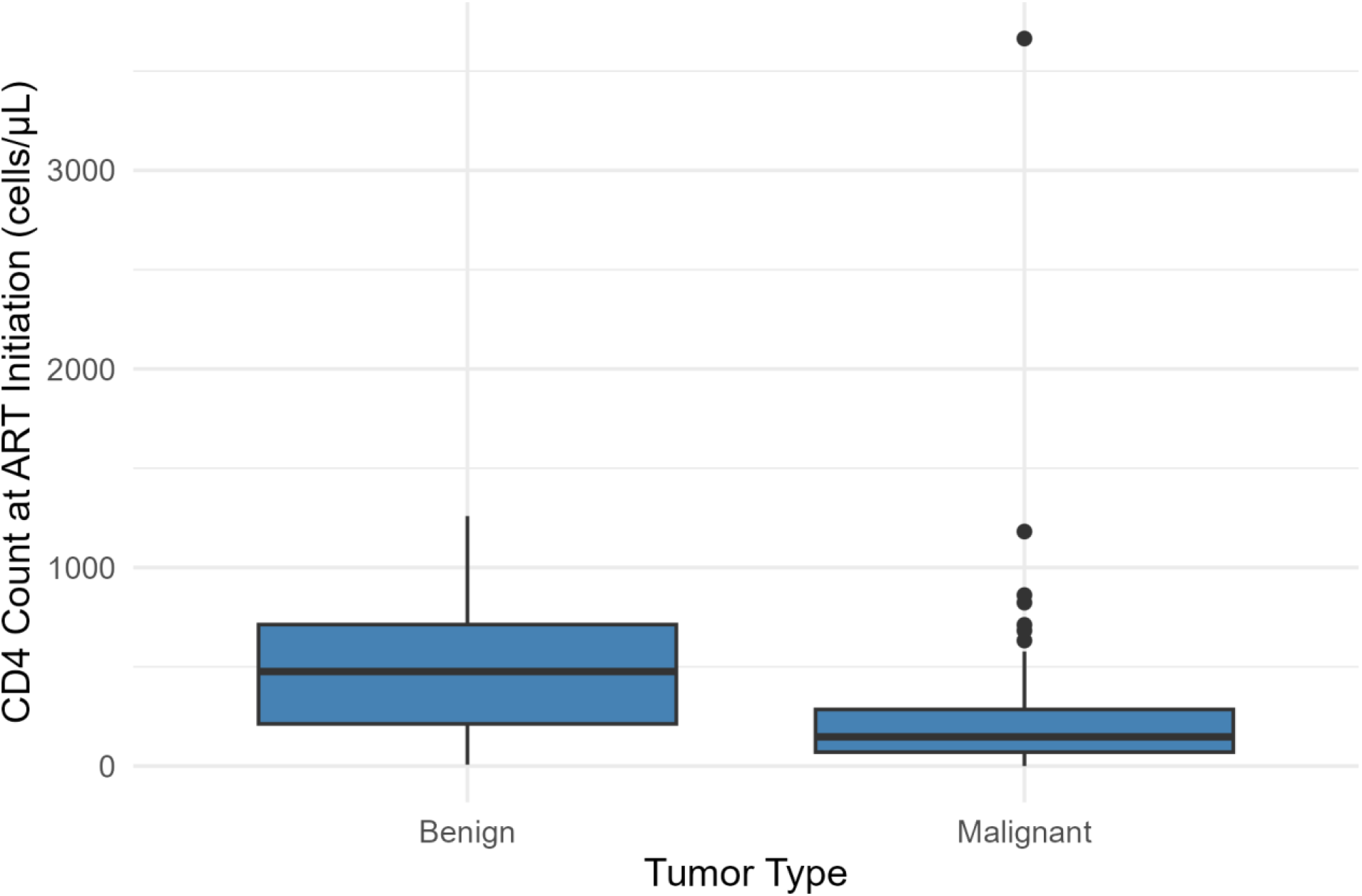
CD4 counts at ART initiation stratified by tumour malignancy status. Box plots display median, interquartile range (IQR), and 1.5×IQR whiskers; individual outliers are shown as points. The malignant group shows a markedly compressed CD4 distribution (median 147 cells/µL) compared with benign tumours (median 476 cells/µL; Mann–Whitney U, p<0.001). N = 93 records with available CD4 data.

**Figure 3.**
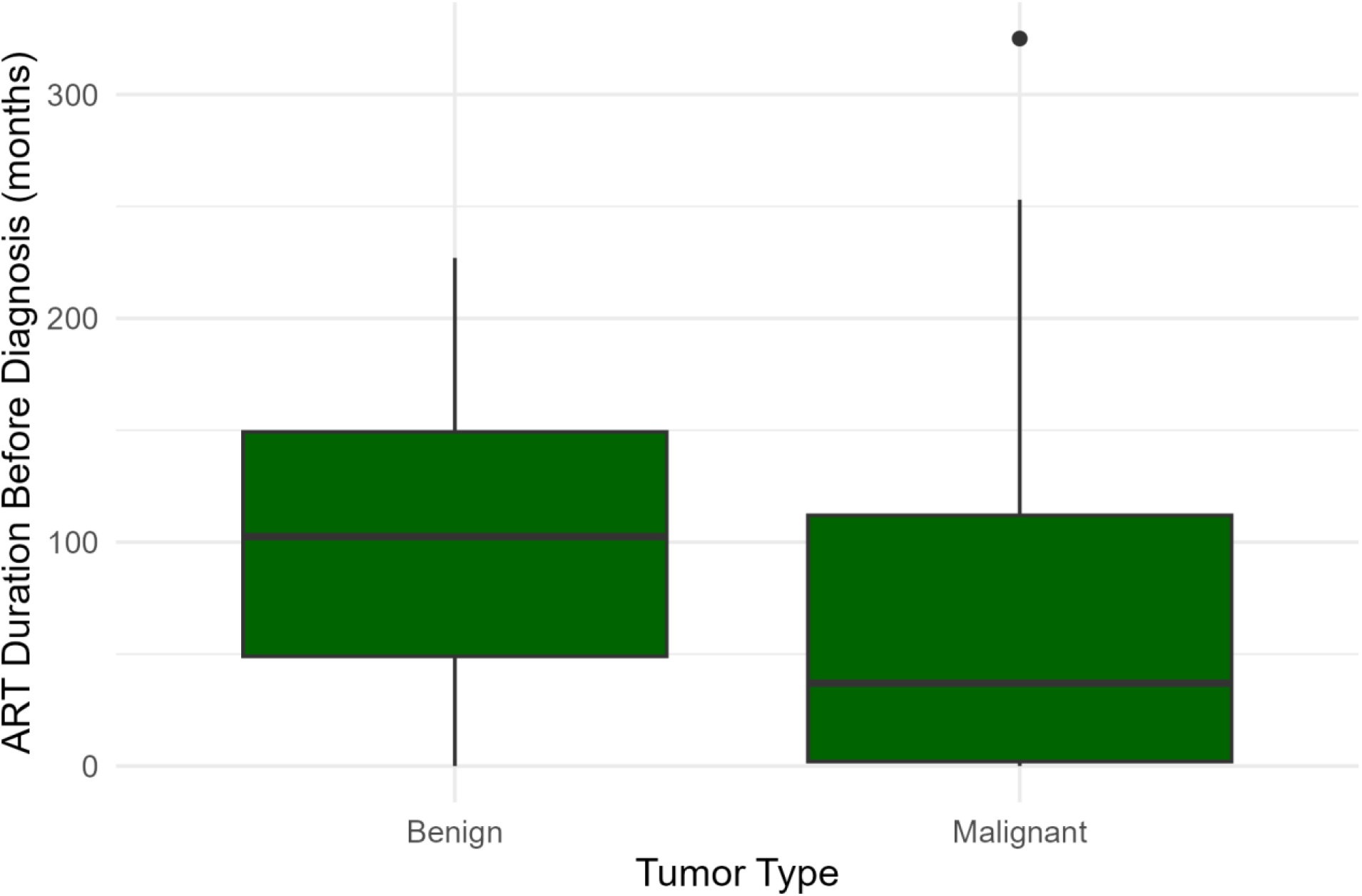
ART duration before tumour diagnosis stratified by tumour malignancy status. Box plots display median, IQR, and 1.5×IQR whiskers; individual outliers shown as points. Malignant tumours were diagnosed after substantially shorter ART exposure (median 37 months) compared with benign tumours (median 102 months; Mann–Whitney U, p<0.001). N = 211 records with available ART duration data.

#### WHO clinical stage at ART initiation

Advanced WHO stage (III/IV) at ART initiation was strongly associated with malignant tumour diagnosis: 39/86 (45.3%) of patients with malignant tumours had advanced stage at ART start, compared with 9/79 (11.4%) of those with benign tumours (χ^2^ *p*=0.002; OR 3.53, 95% CI 1.59–7.84). The gradient was striking among the 25 patients in WHO stage IV at ART initiation, 24 (96.0%) were subsequently diagnosed with a malignant tumour, of whom 20 (83.3%) had KS. This represents a near-complete enrichment of malignancy among the most immunologically compromised patients at ART entry (Figure 1).

#### CD4 count at ART initiation

Among the 93 records with available CD4 data, median pre-ART CD4 was markedly and significantly lower in patients ultimately diagnosed with malignant tumours compared with those with benign tumours (147 vs 476 cells/µL; Mann–Whitney U p<0.001). The categorical analysis reinforced this gradient: 44/71 (62.0%) patients with pre-ART CD4 <200 cells/µL developed a malignant tumour, compared with 8/19 (42.1%) of those with CD4 ≥500 cells/µL (χ^2^ p<0.001) (Figure 2). The high missing data rate for CD4 (57.7%) reflects that many patients transferred in or were enrolled after initial workup at other facilities.

#### ART duration before tumour diagnosis

Median ART duration before tumour diagnosis was substantially shorter for malignant tumours (37 months; IQR 2–112) compared with benign tumours (102 months; IQR 49–149; p<0.001). Malignancy risk was heavily concentrated in the earliest ART period: 54/62 (87.1%) of patients diagnosed within ≤12 months of ART initiation had a malignant tumour, declining to 31/64 (48.4%) among those on ART for >120 months (Figure 3). This temporal gradient supports an interpretation of both unmasking IRIS-associated KS and detection of previously occult malignancies during early clinical engagement.

#### HIV RNA at tumour diagnosis

Among 156 patients with viral load data, 134 (85.9%) were virally suppressed. Although unsuppressed viraemia was numerically associated with a higher malignant proportion (16/22, 72.7% vs 70/134, 52.2%; OR 2.44, 95% CI 0.90–6.61), this difference did not reach statistical significance (Fisher exact *p*=0.105). Critically, the absolute majority of malignant tumours (70/86, 81.4%) occurred in virally suppressed patients, a pattern that underscores the insufficiency of viral suppression alone as a cancer risk stratification tool.

#### ART regimen category

The malignant fraction was nearly identical across regimen classes (DTG-based 49/77 [63.6%]; EFV-based 32/50 [64.0%]; other 33/51 [64.7%]; χ^2^ *p*=0.992), providing no evidence that regimen class itself modifies malignancy risk in this cohort.

### 3.5 Kaposi sarcoma versus other malignancies: immunovirological contrast

Among the 138 malignant tumours, KS patients (n=65) were distinguished from patients with other malignancies (n=73) by substantially lower pre-ART CD4 counts (136 cells/µL, IQR 53–194 vs 269 cells/µL, IQR 128–593; *p*=0.002) and markedly shorter ART duration before tumour diagnosis (4 months, IQR 0–63 vs 86 months, IQR 30–137; p<0.001). KS patients were also significantly more likely to be male (41/65, 63.1% vs 32/73, 43.8%; *p*=0.024) and to have had advanced WHO stage at ART initiation (65.8% vs 29.2% with available data; *p*=0.001). Interestingly, viral suppression rates at the time of tumour diagnosis did not differ between KS and other malignancies (81.3% vs 81.6%; *p*=0.97), reinforcing the point that KS in Uganda frequently arises in virally suppressed but immunologically vulnerable individuals. These findings are summarised in Table 4 and illustrated in Figures 4 and 5.

**Table 4.** Immunovirological comparison of Kaposi sarcoma versus other malignancies among patients with malignant tumours (n = 138).

| Variable | Kaposi Sarcoma<br>(n=65) | Non-KS<br>malignancies (n=73) | p-value | Test |
| --- | --- | --- | --- | --- |
| CD4 at ART initiation,<br>cells/μL, median (IQR) | 136 (53–194) | 269 (128–593) | 0.002 | Mann–Whitney<br>U |
| ART duration before tumour<br>diagnosis, months, median<br>(IQR) | 4 (0–63) | 86 (30–137) | <0.001 | Mann–Whitney<br>U |
| Male sex, n (%) | 41 (63.1) | 32 (43.8) | 0.024 | $\chi^2$ |
| Advanced WHO stage<br>(III/IV) at ART initiation, n<br>(%) | 25/38<sup>a</sup> (65.8) | 14/48<sup>a</sup> (29.2) | 0.001 | $\chi^2$ |
| Virally suppressed at tumour<br>diagnosis, n (%) | 39/48<sup>a</sup> (81.3) | 31/38<sup>a</sup> (81.6) | 0.97 | $\chi^2$ |

<sup>a</sup> Denominator reflects records with non-missing data for the variable. KS, Kaposi sarcoma; IQR, interquartile range.

**Figure 4.**
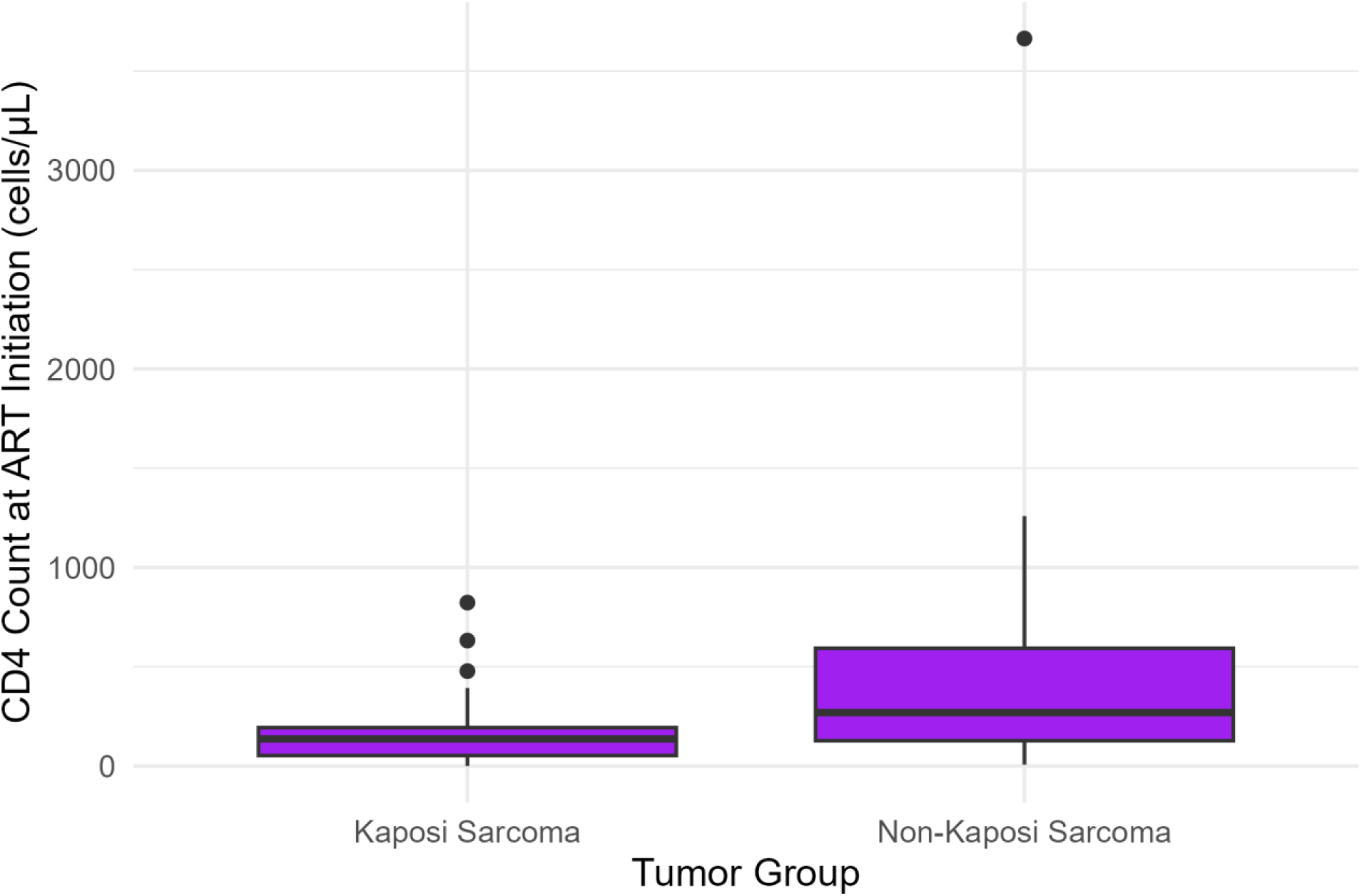
CD4 count at ART initiation in Kaposi sarcoma versus non-KS malignancies. Analysis restricted to malignant tumours (N = 93 with available CD4 data). KS patients had substantially lower pre-ART CD4 counts (median 136 cells/µL, IQR 53–194) than patients with other malignancies (median 269 cells/µL, IQR 128–593; Mann–Whitney U, *p*=0.002).

**Figure 5.**
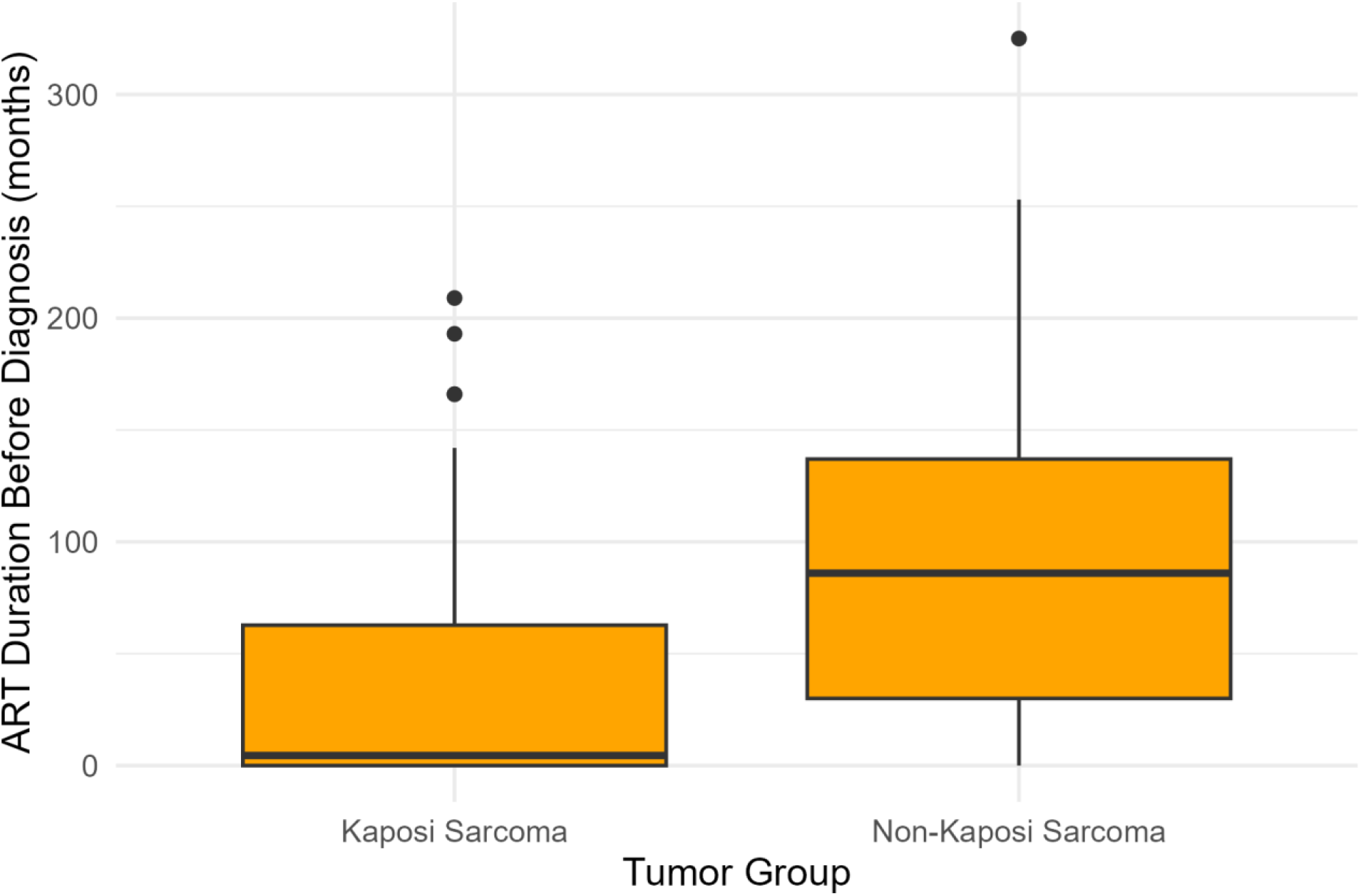
ART duration before tumour diagnosis in Kaposi sarcoma versus non-KS malignancies. Analysis restricted to malignant tumours (N = 131 with available ART duration data). KS patients were diagnosed significantly earlier in the ART course (median 4 months, IQR 0–63) than patients with other malignancies (median 86 months, IQR 30–137; Mann–Whitney U, p<0.001).

### 3.6 Temporal pattern of tumour registrations

Annual tumour registrations peaked in 2018 (n=73), reflecting a data quality improvement programme, then settled to 10–19 records per year between 2020 and 2024, with a modest increase to 18 in 2025. The pre-2020 peak should not be interpreted as a true epidemiological surge in cancer incidence.

## 4. Discussion

This study to our knowledge has provided the first immunovirologically contextualised description of the tumour spectrum among PLHIV at a specialised HIV Centre in Uganda during the transition to DTG-based ART. Six findings deserve emphasis.

### First, KS remains the dominant malignancy despite near-universal viral suppression

KS accounted for 47.1% of all malignant tumours and 29.5% of the entire cohort. This is consistent with sub-regional cancer registry data from Uganda [13,14,18] and broader East African evidence [9,10], but is striking against the declining KS trends observed in high-income settings [3,5]. The biological rationale is compelling: HHV-8 seroprevalence in Ugandan adults exceeds 40% [27,28], HHV-8 reactivation persists in many virally suppressed PLHIV [29,30], and the immune compartments responsible for HHV-8 surveillance, notably CD8 T-cells and natural killer cells recover incompletely following late ART initiation [31]. The finding that 96% of patients in WHO stage IV at ART initiation subsequently developed a malignant tumour, with KS accounting for 83% of those, makes a compelling case for lifelong, viral-load-independent KS surveillance in high-HHV-8-burden settings.

### Second, the virological suppression paradox is firmly established in this cohort

The finding that over 80% of malignant tumours occurred in virally suppressed patients is a pattern now well recognised in high-income cohorts [4,32,33] but rarely demonstrated in routine HIV clinic data from sub-Saharan Africa. The mechanistic implications are substantive: HIV viral suppression does not automatically restore immune competence against oncogenic co-infections, nor does it normalise chronic immune activation, immune senescence, or microbial translocation pathways increasingly implicated in HIV-associated cancer biology [4,6,34]. Operationally, this observation means that viral load status alone is an insufficient criterion for cancer risk stratification in HIV programmes.

### Third, the immunological footprint of late HIV diagnosis is durable

The strong gradient between pre-ART CD4 count (median 147 vs 476 cells/µL in malignant vs benign tumours) and subsequent cancer diagnosis, even after years of suppressive ART, reaffirms that pre-treatment immune damage creates a persistent oncological vulnerability [35,36]. This has direct policy relevance for Uganda’s UNAIDS 95-95-95 target attainment: every avoided late HIV presentation is also a potentially avoided future cancer, justifying sustained investment in community testing and linkage to care.

### Fourth, malignancies cluster in the early ART period with important dual explanations

With 87.1% of tumours diagnosed within 12 months of ART initiation being malignant, this period carries a sharply elevated malignancy burden. Two non-mutually exclusive mechanisms operate: immune reconstitution inflammatory syndrome (IRIS) unmasking of subclinical KS and other cancers [37], and the detection of pre-existing occult malignancies during the intensified clinical engagement that accompanies ART initiation. The clinical implication is that systematic tumour evaluation including careful dermatological examination, lymph node assessment, and structured symptom screening should be embedded within the first-year ART follow-up schedule.

### Fifth, the immunovirological profile of KS is distinctively severe compared with other malignancies

The new KS-versus-non-KS analysis in this study (Table 4) reveals that KS patients had a median pre-ART CD4 of only 136 cells/µL nearly half that of patients with other malignancies and a median ART duration of just 4 months at tumour diagnosis, versus 86 months for other cancers. This demonstrates that KS in this setting arises in a fundamentally different, more immunologically extreme context than other HIV-associated malignancies and should be managed within a distinct high-risk clinical pathway.

### Sixth, the gynaecological burden is dominated by benign disease with a residual cervical cancer signal

Benign uterine leiomyomas constituted the largest single benign tumour category and the second-largest disease group overall, reflecting the feminised HIV epidemic and Mildmay’s integrated women’s health services. The 10 invasive cervical cancers identified against the backdrop of the WHO 2030 cervical cancer elimination strategy and Uganda’s HPV vaccination rollout represent missed prevention opportunities. These findings reinforce calls for systematic, visit-integrated cervical screening for all women living with HIV, as recommended in current guidelines [38].

### Strengths and limitations

The strengths of this study include: use of a longitudinal institutional electronic record that captures HIV treatment history, virological monitoring, and ICD-10-coded tumour diagnoses within a single linked system; a near-decade-long observation window spanning a major ART policy transition; pre-specified analytical decisions; transparent missingness reporting; and a new KS-versus-non-KS comparison that is not typically reported in retrospective HIV-oncology datasets.

The limitations are also substantial and should temper interpretation. The retrospective design, combined with non-trivial missingness for CD4 (57.7%) and viral load (29.1%), introduces potential bias; missingness for CD4 is plausibly ‘not missing at random’ since patients on stable suppressive ART are monitored less frequently, likely creating selection towards sicker patients in the available-CD4 subset. The absence of a denominator of all PLHIV in care at Mildmay precludes calculation of cancer incidence rates. Reliance on ICD-10 coding without universal histological confirmation means some diagnoses may reflect clinical rather than pathological ascertainment. The 2018 registration peak partly reflects a data quality programme rather than true incidence change. Three patients contributed two tumour records each; we analysed by tumour record to preserve cancer spectrum focus and have disclosed this in the Methods. No multivariable regression was performed given the descriptive aim and missing-data structure; all associations presented are hypothesis-generating rather than causal.

### Implications for research and clinical practice

These findings support a programmatic research agenda at Mildmay Research Centre comprising: (i) a prospective HIV-oncology cohort with systematic tumour ascertainment, histological confirmation, and biobanking to allow HHV-8 immune profiling; (ii) a nested study of HHV-8 viral dynamics and cellular immune reconstitution trajectories among virally suppressed PLHIV with and without KS; (iii) integration of HPV-based cervical screening and screen-and-treat into routine HIV care for all women; and (iv) implementation research on the yield, cost, and acceptability of structured cancer symptom screening embedded in early ART follow-up visits. Each priority is operationally feasible at our Centre and directly responsive to donor priorities articulated by the WHO Global Initiative for Cervical Cancer Elimination and the NCI/NIH commitment to global cancer research in PLHIV [38].

## 5. Conclusions

Among PLHIV attending a specialised HIV Centre in Uganda, tumours are predominantly malignant and dominated by Kaposi sarcoma, even against a backdrop of high viral suppression rates. Malignancy clusters among patients who initiated ART late with low CD4 counts and advanced WHO stage and concentrates in the first year of treatment. A new, clinically important finding is that KS arises in a distinctly more immunologically compromised context than other malignancies in this cohort. These data delineate a tractable clinical research and surveillance agenda: lifelong cancer monitoring for all PLHIV regardless of viral load status; structured, systematic tumour evaluation during the early ART period; integration of cervical cancer screening into routine HIV care; and a prospective HIV-oncology cohort to disentangle the residual mechanisms; immune senescence, incomplete reconstitution, oncoviral co-infection, and chronic inflammation that sustain HIV-associated cancer risk in the virally suppressed.

## Data Availability

The de-identified dataset supporting the findings of this study is available from the corresponding author (Joseph Mwaka) upon reasonable request, subject to the data-sharing policies of Mildmay Research Centre, Kampala, Uganda.

## Declarations

### Ethics approval and consent to participate

Approved by the Mildmay Research Ethics Committee (ref: MUREC-2025-808). Individual informed consent waived for routinely collected, de-identified data.

### Availability of data and materials

The de-identified dataset is available from the corresponding author on reasonable request, subject to institutional data-sharing policies.

### Competing interests

The authors declare no competing interests.

### Funding

This work received no dedicated funding.

### Authors’ contributions

Joseph Mwaka conceived the study, oversaw data extraction, led immunovirological interpretation, and drafted the manuscript. Jessica Natalayi conducted all statistical analyses and generated figures. Conrad Sserunjogi curated clinical data and verified ICD-10 coding. All authors interpreted findings, critically revised the manuscript, and approved the final version.

## Acknowledgements

We thank the patients of Mildmay Hospital and the clinical and data teams whose dedicated work made this analysis possible.

## Notes

### Competing Interest Statement

The authors have declared no competing interest.

### Funding Statement

This study received no external funding. It was conducted using routinely collected clinical data from Mildmay Hospital, Kampala, Uganda. The research was carried out through Mildmay Research Centre, the research arm of Mildmay Uganda. No payments or services were received from any third party for any aspect of this work, including study design, data collection, statistical analysis, or manuscript preparation.

### Author Declarations

The Ethics Committee of Mildmay Research Centre, Kampala, Uganda gave ethical approval for this work (reference: MUREC-2025-808). Given the retrospective nature of the study and use of de-identified routine clinical records, the requirement for individual informed consent was waived by the same committee.

